# Estimating the infection fatality ratio of zoonotic avian influenza viruses with pandemic potential using an evolutionary epidemiological model

**DOI:** 10.64898/2026.01.21.26344526

**Authors:** Joshua Mack, Michael Li, Amy Hurford

## Abstract

The risk of zoonotic avian influenza (AIV) infection to humans is challenging to estimate as many human avian influenza virus infections are undetected because infections may be asymptomatic, symptomatic but not tested, and difficult to identify through contact tracing, as human-to-human transmission is rare. We derive equations that consider the evolutionary mechanisms that give rise to pandemics and are parameterized to be consistent with records of past pandemics. We estimate that thousands of human infections with AIVs possessing pandemic potential occur worldwide in an average year. Combining these estimates with H5N1 fatality data, we estimate a historical average infection fatality ratio of 32 (95% uncertainty interval: 9.6– 75) deaths per 10,000 infections. This estimate is comparable to SARS-CoV-2 during the recent pandemic and higher than seasonal human influenza. We estimate that preventing animal-to- human influenza spillovers would delay pandemic emergence by several years. Preventing human infections with AIVs is necessary given the high risk of severe outcomes to individuals and to reduce the risk of pandemics occurring in the future.

## 1. Introduction

The spread of avian influenza viruses (AIVs) in animals poses a risk to humans both individually, due to the risk of zoonotic infections that result in severe outcomes, and collectively, due to the risk that AIVs adapt to transmit effectively in humans and cause a pandemic. Past influenza pandemics have occurred due to reassortment between human influenza viruses and AIVs resulting in strains that spread between humans [1]. Reassortment is the swapping of gene segments between viruses [2] and is a mechanism for the evolution of mammalian-adapted strains of AIVs [3]. Recent AIV outbreaks have occurred in wild birds, poultry, and mammals, with some human infections reported. In 2022, highly pathogenic avian influenza (HPAI) A(H5N1) virus spread across North America and caused mass mortality among wild birds in Canada with tens of thousands of birds reported sick or dead [4,5]. An HPAI A(H5N1) virus outbreak in U.S. poultry and cattle has infected 71 people since March 2024 [6]. The currently circulating H5N1 viruses have not yet adapted to spread well in humans, and of these recent infections there is no evidence of ongoing human-to-human spread [7]. Recent H5N1 virus transmission events among mammals include farmed mink in Europe and marine mammals in South America [8], and mammalian-adapted substitutions have been found in recovered viruses from infected mammals in Canada [3].

The transmissibility of zoonotic AIVs from animals-to-humans, human-to-human, and the pathogenicity of AIV infections in humans depends on characteristics of the influenza virus and host factors. Influenza virus virions have two surface envelope proteins: hemagglutinin (HA) and neuraminidase (NA). HA binds to sialic acid whose presence, location and structure vary between species which is a major reason for host specificity. There are 18 HA subtypes and 9 NA subtypes with H5 and H7 being able to mutate from low pathogenic to highly pathogenic [9,10] . Zoonotic spillover refers to the spread of a pathogen from animals to humans [11]. For zoonotic spillover events, the introduced strain typically cannot yet spread between humans but may acquire adaptations through reassortment and/or mutation that enable human-to-human transmission and ultimately lead to a pandemic [12,13].

HPAI A(H5N1) virus of the A/goose/Guangdong/1/1996 lineage emerged in Asia in 1996 and since 2003, 986 human cases have been reported around the world with 473 deaths, corresponding to a case fatality ratio of 48% [14,15]. Yet the case fatality ratio (the number of deaths per case) likely does not describe the risk associated with zoonotic AIV infections, as only some infections are reported as cases, and many human infections are undetected as they may be asymptomatic or symptomatic but not tested. In addition, as there are few secondary infections in humans [16], contact tracing to detect infections is usually not possible. By combining observations describing the occurrence of pandemics in the past with a model that describes the epidemiological and evolutionary processes that occur when an animal strain of influenza adapts to cause a pandemic, we can estimate the annual number of human zoonotic AIV infections associated with pandemic emergence. The infection fatality ratio (IFR) is the number of deaths per infection [17] and more accurately quantifies the risk of a severe outcome occurring among infected individuals. We use our approach to estimate the IFR associated with zoonotic AIV infections with pandemic potential, using model-estimated infections as the denominator and observed deaths from human HPAI A(H5N1) infections as the numerator. Therefore, our IFR estimate should be interpreted as a historical average severity estimate for pandemic-potential zoonotic AIV infections, informed by available H5N1 fatality data.

The benefit of preventing human infections with zoonotic AIVs is not only to the individual, as can be quantified by the IFR, but also to reduce the chance that evolutionary change occurs resulting in a pandemic. We use our modelling approach to estimate the effect of preventing human infections with AIVs on delaying the time until the next pandemic. Previous studies have modelled the transmission of HPAI A(H5N1) viruses between birds and mammals [18,19], H5N1 virus spillover to humans [20], and the human-to-human transmissibility of H5N1 virus using outbreak data [21]. Our approach overcomes the challenge of limited surveillance to detect asymptomatic human AIV infections. The objectives of this study are to estimate the annual number of human infections caused by zoonotic AIVs with pandemic potential, estimate the associated IFR using HPAI A(H5N1) fatality data, and determine how preventing zoonotic spillovers of AIV to humans would delay a future pandemic.

## 2. Methods

### (a) Model formulation

To develop the model to estimate the annual number of human AIV infections from the historical rate of pandemics, we adapted the approach of Day et al., 2006 [13] who estimated the minimum annual number of human AIV infections. Throughout this manuscript, we use the term AIV to refer to zoonotic avian influenza viruses with pandemic potential, unless otherwise specified.

Instead of using analytical approximations to calculate the *minimum* annual number of human AIV infections as was done in [13], we used numerical methods to estimate the *mean* annual number of human AIV infections. Following [13], we developed a stochastic model that describes the epidemiological and evolutionary processes that give rise to an influenza pandemic of zoonotic origin. The quantities used to predict the annual probability of a pandemic are the number of humans infected by zoonotic spillovers, the transmissibility of AIV in humans, and the probability of, and epidemiological characteristics that arise from, evolutionary change. All R scripts and output files required to reproduce the analyses are publicly available (see the Data Availability Statement).

We use a multitype branching process to model human infections with a genotype, *i*, of an AIV. Given *σ* different AIV genotypes, each that are circulating in non-human animal populations with frequencies, *ρ ρ*_2_,…, *ρ_σ_* then the probability mass function for the number of human infections with AIV is,

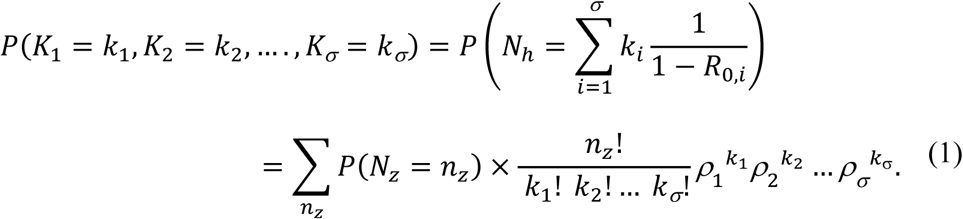

The zoonotic AIVs may evolve to infect other animals besides birds, and we let *n_z_* be the annual number of zoonotic spillovers (i.e., AIV infections that spread from animals to humans), where *N_z_* is a random variable describing this quantity. We define *R*_0,*i*_ as the basic reproduction number of an AIV of genotype *i* introduced to humans prior to any adaptation. This is the number of secondary human infections generated by a human infected with an AIV in a wholly susceptible human population before any evolutionary change occurs during any of the subsequent human infections. This basic reproduction number, *R*_0,*i*_, is required to be less than 1 because it is assumed that the AIV genotype that spills over to humans is not yet adapted for human-to-human spread [13,22].

The term 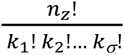 is the possible combinations of numbers of genotypes that make up all the *n_z_* zoonotic spillovers, which is multiplied by the probability of these combinations occurring, 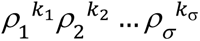, and together these two terms describe the probability mass function of a multinomial distribution, conditioned on a given number of zoonotic spillovers, *n*_z_. The multiplication by the probability of *n*_z_ zoonotic spillovers, *P*(*N_z_* = *n_z_*), and the sum over *n*_z_ reflects the law of total probability. The righthand side of equation (1) is the probability that the number of spillovers for each of the genotypes is *K*_1_ = *k*_1_, *K*_2_ = *k*_2_,…, *Kσ* = *kσ*, and this is equated to the probability mass function for the number of human AIV infections, *P*(*N*_*h*_ = *n*_*h*_), that occur for this number of spillovers of each genotype. Specifically, the genotype *i* is expected to generate 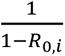 human infections [23], there are *k_i_* instances of zoonotic spillover due to genotype *i*, and the total number of human infections is the sum across all genotypes.

There are different subtypes and clades among zoonotic AIVs and these aspects of influenza virology are represented in our modelling as we consider AIV genotypes, *i*. Genotypes that have characteristics, such as high transmissibility in humans, may be more likely to be of a particular subtype or clade, but our modelling represents only the diversity in the epidemiological characteristics without associating such characteristics to a particular subtype.

We assume the number of zoonotic spillovers can be modelled by a probability mass function *P*(*N_z_* = *n_z_*) with a mean equal to *n̄_z_*. Calculating the expected value gives the mean number of human infections with AIV that occur annually, *n̄*_*h*_, as,

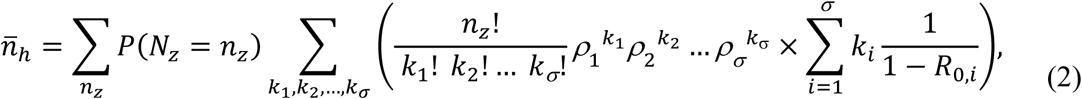

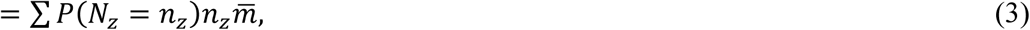

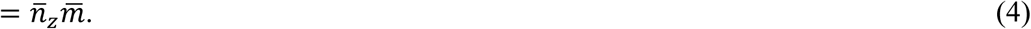

As such, the mean annual number of human infections includes both the zoonotic spillovers and any subsequent human-to-human transmission of AIV. Equation (4) relates the quantity that we aim to estimate, *n̄*_*h*_, to the mean number of zoonotic spillovers of AIV, *n̄_z_*, which we will show can be estimated from the historical rate of pandemics. The simplification that occurs in equation (3) is due to the expected value of the function 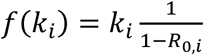 for a multinomial distribution, which is 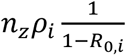. We define 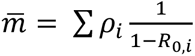 as the mean number of human secondary infections in a wholly susceptible population where the mean is calculated across all genotypes, *i*. The simplification that occurs in equation (4) is due to substituting *n̄_z_*, the expected value of *N_z_* into equation (3).

The probability of a pandemic occurring in any year can be modelled as a random variable *Λ* with a probability density that is,

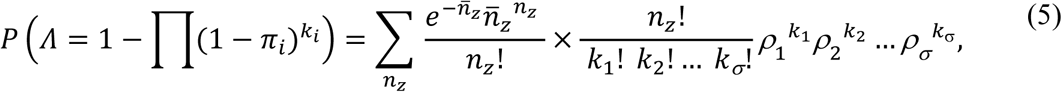

where *πi* is the probability that an infection with the genotype *i* causes a pandemic and *Λ* is equated to the event that at least one of the AIV spillovers results in a pandemic. Data describing the interpandemic periods for seven pandemics that occurred over the last 245 years are used to estimate the annual probability of a pandemic. The annual probability of a pandemic (which can be estimated from data as described in (b)) is equated to the expected value of equation (5), which is

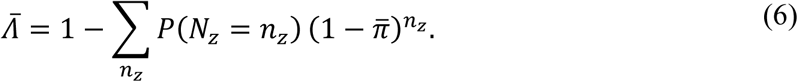

The mean probability that a human infection with the AIV genotype *i* results in a pandemic, *π̄*, depends on the probability that a pandemic-capable strain emerges during a single human AIV infection, *a_i_*, and the probability that the pandemic-capable strain avoids stochastic extinction and causes a pandemic, *P_i_*. This is

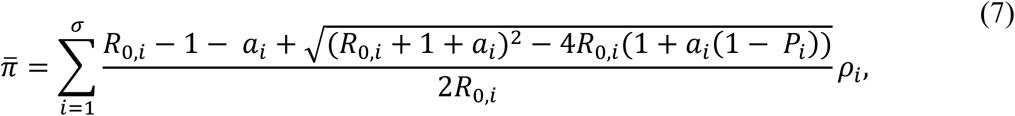

which is equation 2.1a in [13]. Under the branching process model described above, *P_i_* is determined by the basic reproduction number of the pandemic-capable strain, *R*^∗^*_i_*, defined as the expected number of secondary human infections generated by a typical infected individual in a wholly susceptible population. Specifically, *P_i_* = 1 − 1/*R*^∗^*_i_* [12,13], where *R*^∗^*_i_* > 1, consistent with the definition of a pandemic-capable strain.

Our derivation in this section is similar to [13] except that we focus on the mean, rather than the minimum, number of human AIV infections (where the latter was the focus of [13]). As such, we do not use any of the approximations that assume the number of zoonotic spillovers, *n̄_z_π̄*, and the genotypic variance are small.

### (b) Generating genotypes and estimating the mean number of zoonotic spillovers from data

We used Latin hypercube sampling (LHS) [24] to generate genotypes indexed by *i* = 1 to *σ* = 1,000 by sampling from the distributions described in Table 1 (see section A.1 in S1 Appendix for parameter distributions). All influenza A subtypes have the potential to cause the emergence of pandemic strains through reassortment [25] and/or mutation, so we do not specify any particular HA subtype for the sampled genotypes.

**Table 1.**
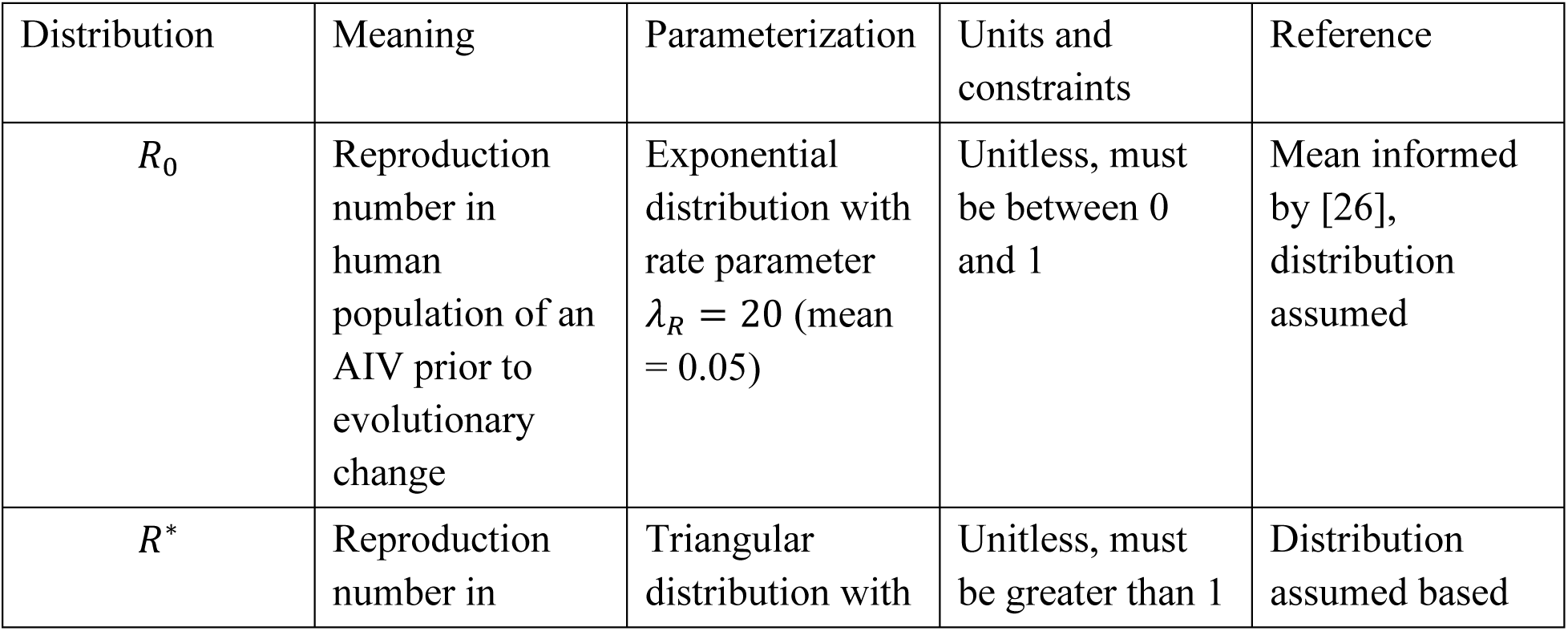
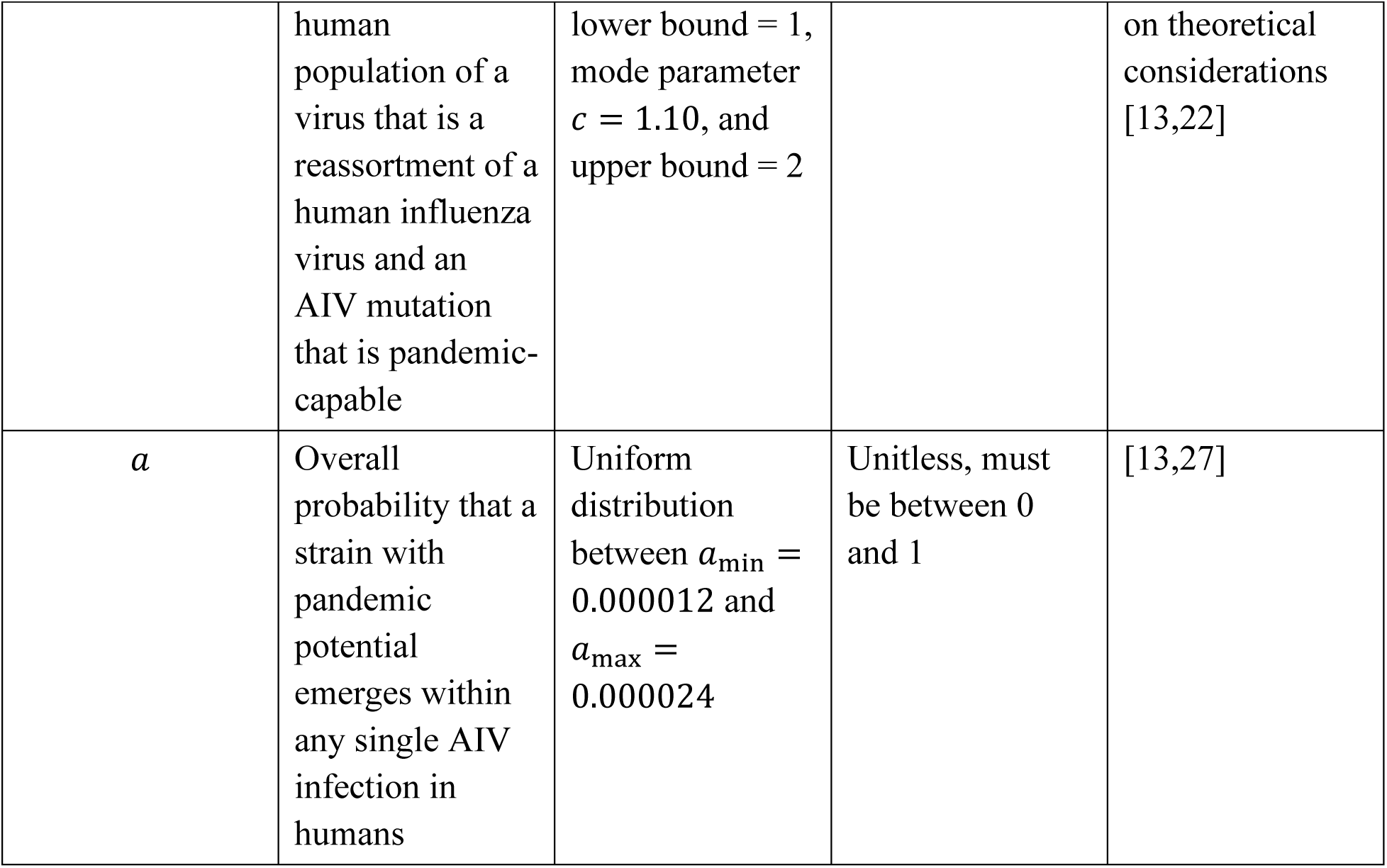
Distributions used for LHS to generate genotypes.

The distributions used to generate genotypes were selected based on available empirical estimates where available and theoretical considerations where empirical data were lacking. For *R*_0_, the reproduction number of an AIV in the human population prior to evolutionary change, we assumed an exponential distribution with rate parameter *λ_R_* = 20, corresponding to a mean of 1/*λ_R_* = 0.05. The exponential distribution was chosen to represent the predominance of AIV genotypes with limited human-to-human transmission potential while allowing for rare, more transmissible genotypes. The mean parameter was informed by published estimates of the reproduction number of HPAI A(H5N1) infections in humans. Recent estimates from the ongoing U.S. H5N1 outbreak suggest a median *R*_0_ of approximately 0.04–0.05, consistent with previous estimates of H5N1 transmissibility from outbreaks in Indonesia, Vietnam, and the broader Asian region [26].

No empirical estimates exist for *R*^∗^, the adapted-virus reproduction number. Therefore, *R*^∗^ was assigned a triangular distribution with a lower bound of 1, an upper bound of 2, and a mode parameter *c* = 1.10 based on theoretical considerations. The mode was placed slightly above the epidemic threshold to represent the assumption that newly emerging pandemic viruses initially exhibit limited but sustained transmissibility [13,22]. The upper bound of 2 was selected to allow for the possibility of moderately transmissible adapted viruses while avoiding assigning substantial probability to highly transmissible viruses that are less consistent with the emergence process represented in the model.

The probability of pandemic-capable evolution, *a*, was based on the estimates of [13]. Following the assumptions of [27], the probability of co-infection between zoonotic AIV and seasonal human influenza was estimated from a seasonal influenza attack rate of 10–20%, a 12-week influenza season, and a one-day opportunity for co-infection during AIV infection. These assumptions were combined with an assumed reassortment probability in [13] to obtain an estimated probability of pandemic-capable evolution ranging from *a*_min_ = 0.000012 to *a*_max_ = 0.000024 per human AIV infection. We represented this uncertainty using a uniform distribution.

Data describing the interpandemic periods for seven influenza pandemics of zoonotic origin that occurred during the past 245 years were used to estimate the historical rate of pandemic emergence (Table 2 and Fig 1). We modelled interpandemic waiting times using a gamma distribution with shape parameter *α* and scale parameter *θ*, such that the mean waiting time was *δ* = *αθ*. The exponential distribution is a special case of the gamma distribution in which *α* = 1, corresponding to waiting times generated by a homogeneous Poisson process. However, the gamma distribution provides additional flexibility by allowing the variance of waiting times to differ from the mean, thereby accommodating overdispersion or underdispersion in historical interpandemic intervals relative to a Poisson process.

**Fig 1.**
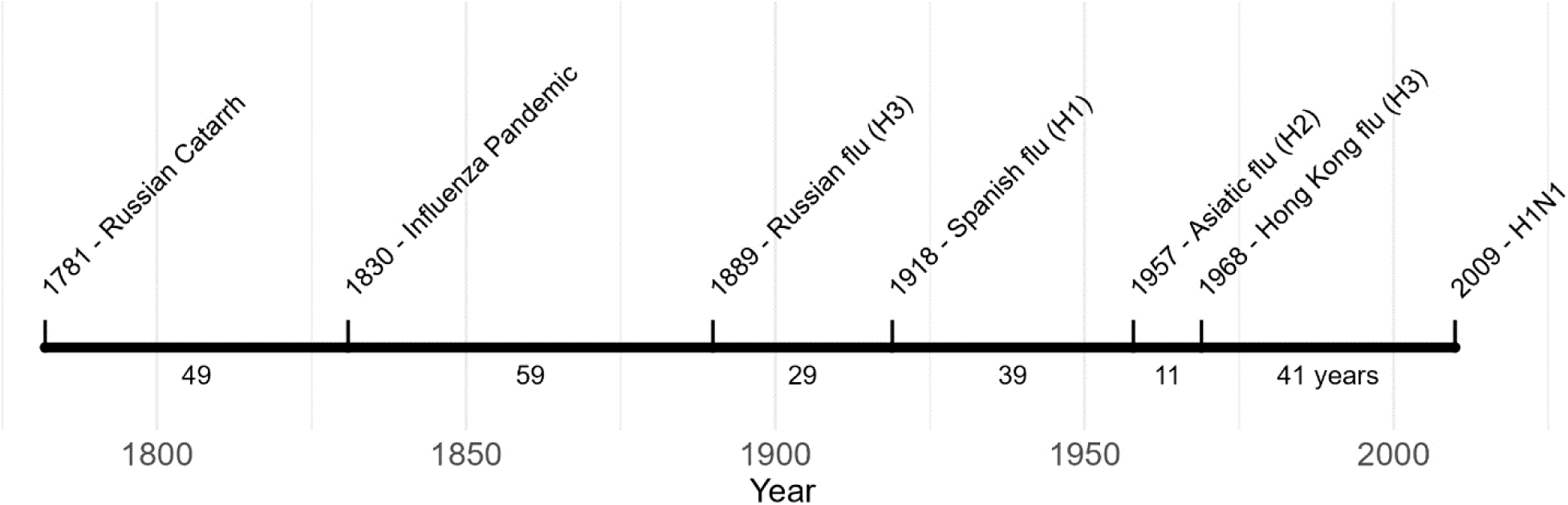
Timeline of influenza pandemics that occurred over the last two centuries with influenza subtypes (if known) and interpandemic period lengths [28–31].

**Table 2.**
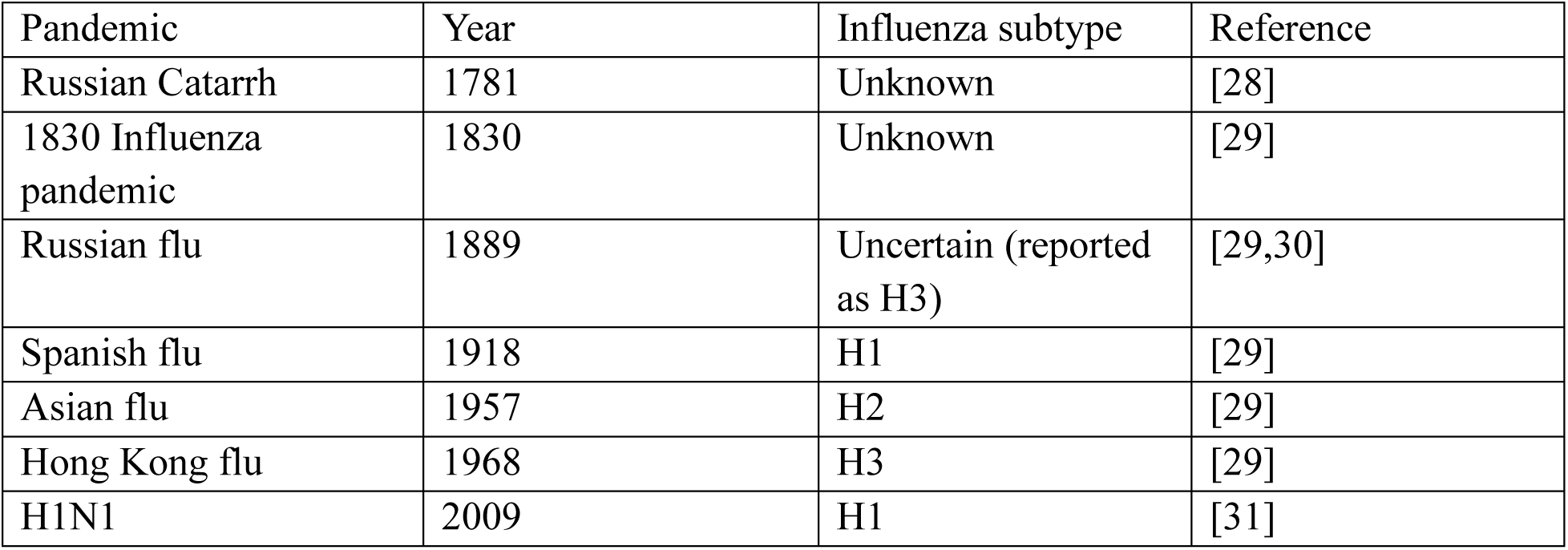
Influenza pandemics of zoonotic origin (data used to fit gamma distribution)

The fitted mean interpandemic period, *δ*, was converted to an annual pandemic probability by assuming that the historical average pandemic rate is the reciprocal of the mean waiting time: *Λ*^ˉ^ = 1/*δ*. This estimated annual pandemic probability was equated with the expected value of the model pandemic probability in equation (6). The gamma distribution was fitted by maximum likelihood using the bbmle package in R [32]. The ongoing interval following the 2009 pandemic was excluded from the likelihood calculation because it represents a right-censored observation rather than a completed waiting time. Because the virological origin of the 1889 pandemic remains uncertain [30] and the 2009 H1N1 pandemic resulted from a swine-origin influenza virus rather than direct AIV spillover [31], sensitivity analyses were conducted excluding the 1889 pandemic, the 2009 pandemic, or both (S1 Appendix A.3).

It has been argued that pandemics are becoming more frequent [33]. As such, we hypothesized that the mean of the interpandemic period is affected by the date of past pandemics, i.e., whereby more recent pandemics may have occurred more frequently. To do this, we let *δ* = *δ*_0_ + *δ*_1_*x_d_* where *δ*_0_ and *δ*_1_ are parameters for the intercept and slope of the relationship between the mean interpandemic period and *x_d_* is the number of years since 1781 (the date of the first pandemic considered). We calculated the likelihood ratio for the model with an effect of recency compared to a model without an effect of recency.

We sampled from our fitted gamma distribution to generate estimates of *Λ̄_i_*. The sampling approach incorporates uncertainty in the interpandemic period, some of which is because there are only a few influenza pandemics reported that occurred during the last few centuries. Our estimation approach does not explicitly consider influenza subtypes because relatively few pandemics have occurred in the past, such that it is not possible to estimate the annual probability of a pandemic due to a particular subtype.

### (c) Calculating the distribution of the mean annual number of human AIV infections

We calculated the distribution of the mean annual number of human AIV infections by completing *σ* = 1,000 samples of *R*_0,*i*_, *P_i_* (calculated as *P_i_* = 1 − 1/*R*^∗^), *a_i_*, and *Λ̄_i_*. This sampling occurred using the lhs package in R. For all the sampled values of *R*_0,*i*_, *P_i_*, and *a_i_*, we calculated *π̄* using equation (7), where the frequency of each genotype is *ρi* = 1/*σ* for all *i*. We then calculated *n̄_z_*_,*i*_ by equating the sampled value of *Λ̄_i_* with *Λ̄* in equation (6) and solving for *n̄_z_*, which is the only unknown value (and for this specific realization of the sampling is referred to as *n̄_z_*_,*i*_). The value of *n̄_z_*_,*i*_ was then substituted for *n̄_z_* in equation (4) to estimate one of the values of *n̄*_*h*,*i*_, where all these values together comprise our estimated distribution for the mean annual number of human infections with AIVs.

### (d) Calculating the IFR

We estimated the IFR for zoonotic AIV infections with pandemic potential by combining our estimate of the annual number of human infections with AIVs with historical fatality data from human HPAI A(H5N1) infections. We calculated the annual number of human AIV-related deaths as the average number of reported human deaths due to H5N1 over the previous 23 years (473 reported deaths [15]), corresponding to 20.6 deaths per year. The IFR was calculated as the annual number of deaths divided by the mean annual number of human infections with AIVs estimated in (c) and was expressed as a percentage. Because H5N1 accounts for most documented fatal human zoonotic AIV infections, we used reported H5N1-associated deaths as a proxy for fatality among pandemic-capable AIV infections. Accordingly, the resulting IFR should be interpreted as an approximate estimate for this broader class of viruses, informed primarily by H5N1-associated fatality, rather than as an H5N1-specific IFR.

### (e) Calculating the impact of preventing zoonotic spillovers

We also investigated the impact of control measures that reduce the mean number of zoonotic AIV spillovers to humans by a fraction, *χ*. We modified equation (6) to determine how these control measures would affect the annual probability of a pandemic as,

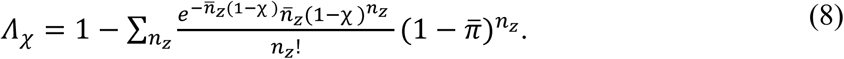

### (f) Comparison to previously estimated minimum annual spillovers

We completed an analysis to compare our results with the minimum annual number of zoonotic spillovers implied by the historical interpandemic period as estimated in [13]. For our comparison, we graphed the range of the minimum estimates of the annual zoonotic spillovers using equation (2.1b) from [13], which is,

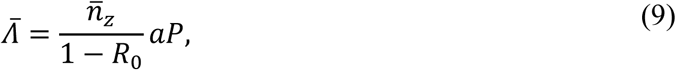

in our notation. As is assumed in Figure 2 of [13], we set *a*_min_ = 0.000012 and *a*_max_ = 0.000024, the minimum and maximum values of *Λ̄* to 0.007 and 0.076, respectively, and fixed *P* = 1. Using this approach, we produced a range for the minimum annual number of zoonotic spillovers as was done in [13].

**Fig. 2.**
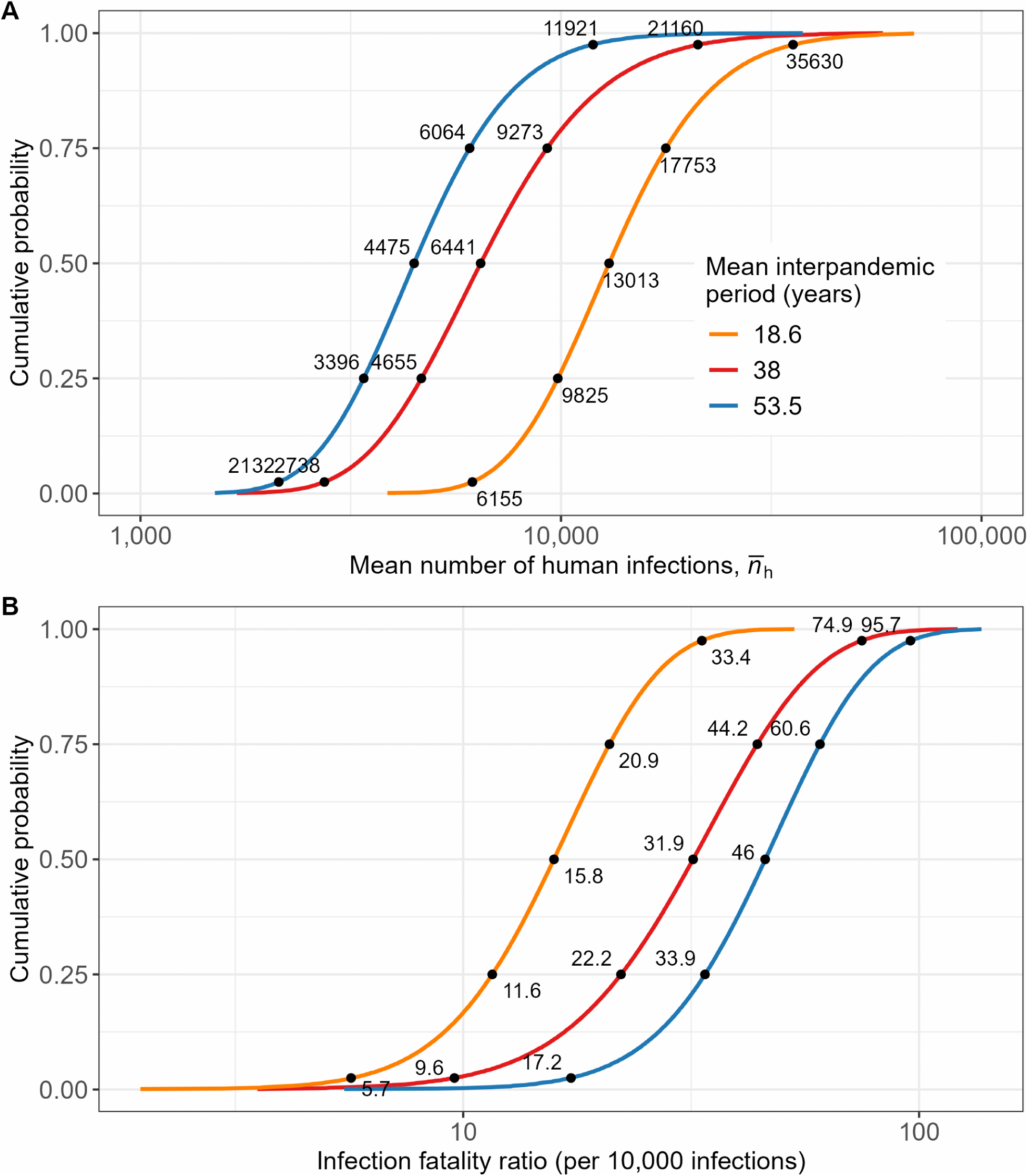
(A) Estimated mean annual number of human infections with AIVs. The baseline model assumes a constant mean interpandemic period of 38 years (red line). Under the time-varying interpandemic period model, the estimated mean interpandemic period was 53.5 years in 1781 (blue line) and 18.6 years in 2026 (orange line). Because the distribution of estimated mean annual human AIV infections is highly right-skewed, estimates are shown on a logarithmic (base 10) scale. (B) Corresponding IFRs for zoonotic AIV infections with pandemic potential, estimated using H5N1-associated deaths as a proxy for AIV- associated fatality.

To estimate an analogous minimum number of zoonotic spillovers using our methodology, we set *P* = 1 for all *i*, set *R*_0_ to each of 0, 0.2, 0.4, 0.6 and 0.8, and sampled from the distributions for the other parameters as described in (b) and (c). Using this approach, we produced a distribution for the minimum annual number of zoonotic spillovers, which we compared to the range that was calculated in [13].

### (g) Sensitivity analyses

We assessed the robustness of the estimated annual number of human AIV infections to key modelling assumptions. One-way sensitivity analyses varied (i) the distributions of *R*_0_, *R*^∗^, and the probability of pandemic-capable evolution, *a*; (ii) assumptions regarding genotype frequencies; (iii) the historical pandemic dataset used to estimate the interpandemic interval; and (iv) the completeness of reporting of human AIV deaths. In addition, we performed a mechanistic boundary analysis in which extreme combinations of parameter values within the assumed ranges were evaluated to determine the maximum and minimum plausible estimates of the mean probability that an AIV infection results in a pandemic, *π̄*, the inferred number of zoonotic spillovers, *n̄_z_*, the mean number of human infections generated per zoonotic spillover, *m̄*, annual human infections, *n̄*_*h*_, and the corresponding IFR. Full parameter specifications and additional results are provided in S1 Appendix A.3.

## 3. Results

We fitted the gamma distribution to the interpandemic periods of past pandemics (see Fig A2 in S1 Appendix) and estimated the mean interpandemic period as *δ* = 38 (95% confidence interval: 25–62) years with *θ* = 8.9 (95% confidence interval: 2.8–28.4). We also fitted a gamma distribution with a mean that depends on the start date of the past pandemics and estimated *δ*_0_ = 53.5 (95% confidence interval: 28.3–102.6) years, *δ*_1_ = −0.14 (75% confidence interval: -0.27– 0.029; standard error: 0.10) years per year, and *θ* = 9.2 (95% confidence interval: 2.7–31.5) (see section A.2 in S1 Appendix for additional details). A likelihood ratio test comparing the nested models with and without an effect of pandemic recency on the mean interpandemic period found no statistically significant improvement in model fit when the recency effect was included (likelihood ratio test: *χ*^2^ = 1.99, *p* = 0.158). While the effect of recency on the interpandemic period is not clear, other studies argue that pandemics are becoming more frequent [33], so we considered results that incorporate this effect in our subsequent analyses.

We sampled from the fitted gamma distribution, generated genotypes as described in (c), and evaluated equation (5) to estimate the distribution of the mean annual number of human AIV infections. The median estimated mean annual number of human AIV infections was 6,441 (95% uncertainty interval: 2,738–21,160) infections per year under the model assuming no temporal trend in the interpandemic period (Fig 2A; red line). This corresponds to an IFR of 0.32% (95% uncertainty interval: 0.096%–0.75%), equivalent to 32 deaths per 10,000 infections (Fig 2B). For reference, the IFR of SARS-CoV-2 in 2020-2023 was estimated as 16 deaths per 10,000 infections [34] and 2.9–6.5 deaths per 10,000 infections for seasonal influenza [35] (Fig 3). These estimates are implied by our calculation of the number of human AIV infections based on the assumed evolutionary and epidemiological process, and the interpandemic periods of the past seven influenza pandemics. The distribution of annual human AIV infections is highly right-skewed, which means that a very large number of human infections with AIVs (*n̄*_*h*_ > 20,000) is possible, although with low probability (< 0.025). The median value of the simulated *n̄*_*h*_ distribution was 6,441 infections per year, which is much lower than the midpoint of the interval between the 2.5^th^ and 97.5^th^ percentiles.

**Fig 3.**
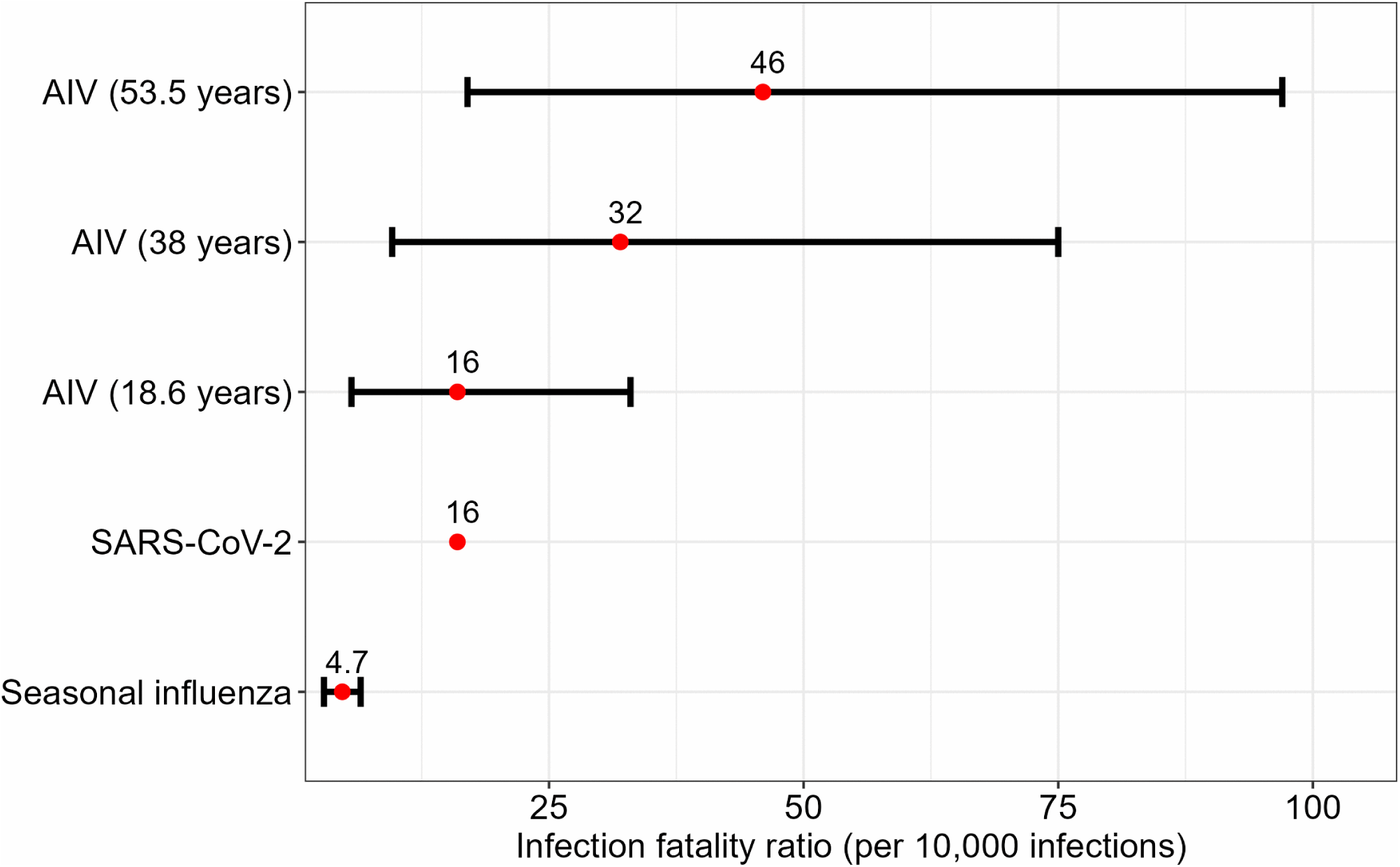
Estimated IFRs (deaths per 10,000 infections) for zoonotic AIV infections with pandemic potential under different mean interpandemic period scenarios, using H5N1- associated deaths as a proxy for AIV-associated fatality, compared with SARS-CoV-2 and seasonal influenza [34,35].

We also calculated how our estimated number of human AIV infections would be different if pandemics are becoming more frequent. The earliest pandemic that we considered occurred in 1781 (Table 2 and Fig 1) and from our statistical analysis, we estimated the mean time to the next pandemic in 1830 to be 53.5 years. For an interpandemic period of 53.5 years, the median estimated mean annual number of human AIV infections was 4,475 (95% uncertainty interval: 2,132–11,921 infections per year; Fig 2A, blue line). This corresponds to an IFR of 46 (95% uncertainty interval: 17–96) deaths per 10,000 infections (Fig 2B). For the year 2026, we estimated the mean interpandemic period to be 18.6 years. Under this scenario, the median estimated mean annual number of human AIV infections was 13,013 (95% uncertainty interval: 6,155–35,630 infections per year; Fig 2A, orange line). This corresponds to an IFR of 16 (95% uncertainty interval: 5.6–33) deaths per 10,000 infections (Fig 2B). Details describing how we calculated the interpandemic period for 1781 and 2026 are provided in section A.2 of S1 Appendix.

We used our parameter estimates and sampling approach to understand the impact of preventing zoonotic spillovers on the annual risk of a pandemic as described by equation (8). We estimated that preventing 5%, 20% and 50% of animal-human AIV spillovers would delay pandemic emergence by an average of 2, 9.4 and 37.5 years respectively (Fig 4). As the annual probability of a pandemic occurring is subject to a high level of uncertainty (as modelled by equation 8), there is some chance, often which is small, that a pandemic can occur sooner than the mean time to the next pandemic when there is no prevention of zoonotic spillovers. Our estimates of the effect of preventing zoonotic spillovers on the number of years the next pandemic would be delayed are subject to a high level of uncertainty (Fig 4).

**Fig 4.**
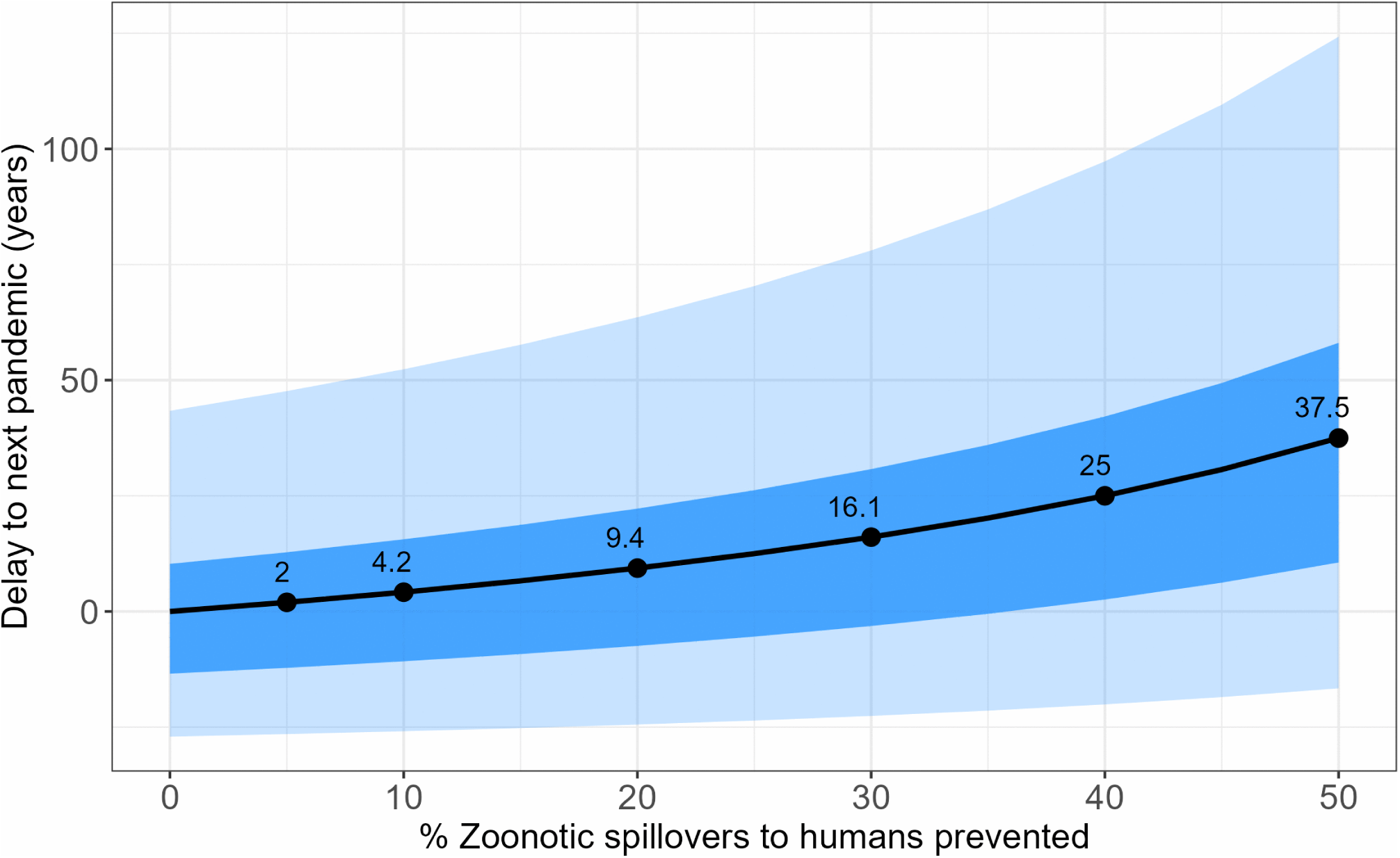
The mean (solid line) number of years that the next pandemic is delayed when a percentage of animal-human AIV spillovers are prevented, with the dark shading representing the interquartile range and the light shading representing the 95% prediction interval. Negative values indicate that the next pandemic is estimated to occur earlier than the absence of spillover prevention; the probability of such outcomes is generally low.

We based our methodology in (a) on [13] but deviated in our approach as we aimed to find the distribution of the mean annual number of human AIV infections rather than the minimum number. We found that our estimates of the minimum annual number of zoonotic AIV spillovers, *n̄_z_* are consistent with [13] (Fig 5A). Our estimates of the mean annual number of zoonotic AIV spillovers, that sample from the distribution for *P,* are higher (Fig 5B) than both the minimum estimates of [13] (grey shading) and our own minimum estimates (Fig 5A).

**Fig 5.**
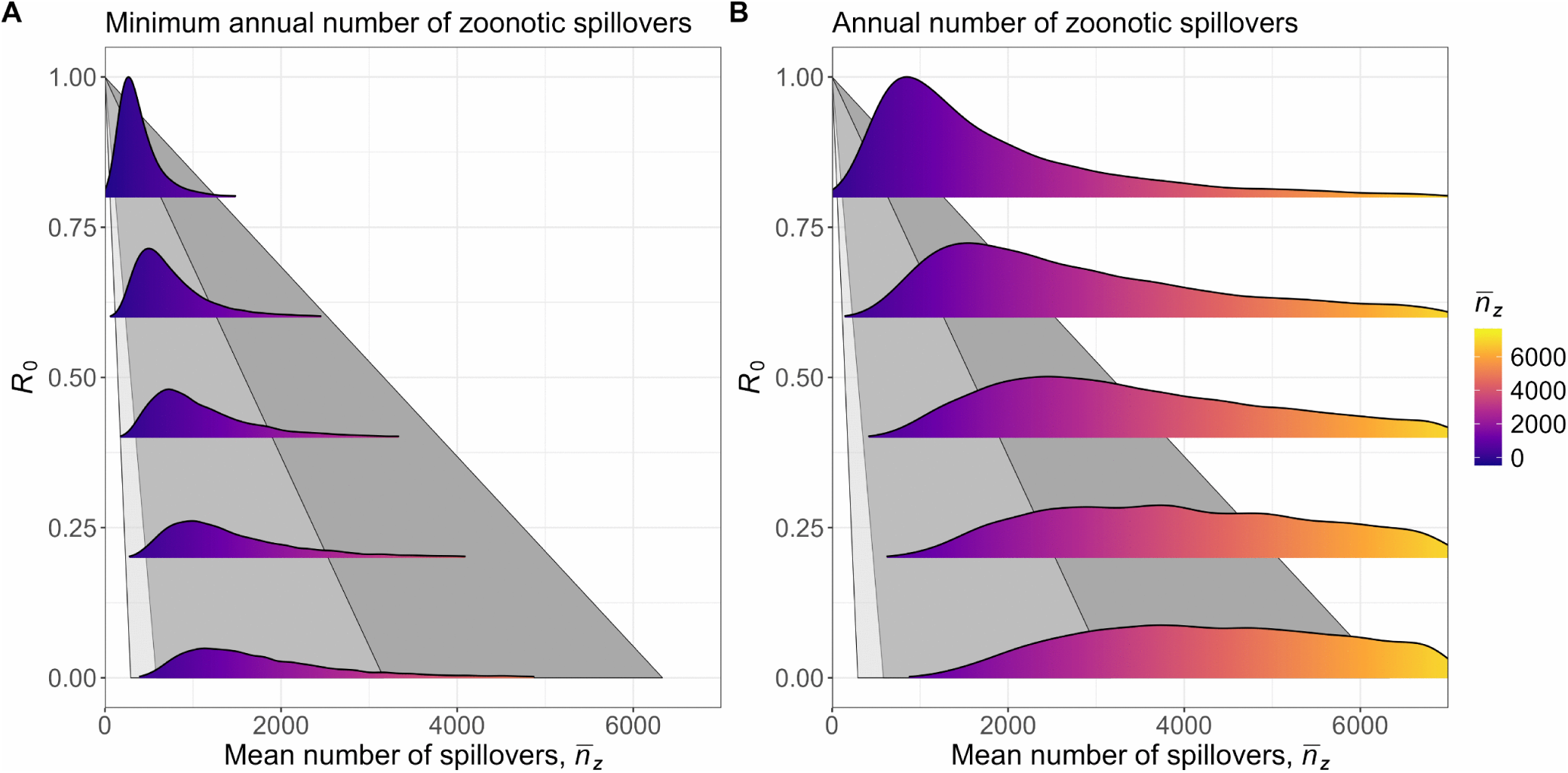
(A) Estimated distribution of the minimum annual number of zoonotic spillovers of AIV (purple shaded curve). (B) Estimated distribution of the mean annual number of zoonotic spillovers (purple-yellow shaded curves). Grey shaded areas reproduce the results from Figure 2 in [13] and span the 95% support interval of the probability of a pandemic occurring in any given year. Light grey shading is an attack rate of 20% and dark grey shading is an attack rate of 10%.

Sensitivity analyses showed that the estimated annual number of human AIV infections had limited sensitivity to plausible variation in assumptions regarding *R*_0_, *R*^∗^, and genotype weighting. In contrast, the estimates were most sensitive to the assumed probability of pandemic-capable evolution, *a*; halving the lower and upper bounds of the assumed range approximately doubled the estimated annual number of infections, whereas doubling the bounds approximately halved the estimate. Sensitivity analyses of the historical pandemic dataset indicated moderate sensitivity to the inclusion of individual historical pandemic events, reflecting the small number of pandemics available to estimate the interpandemic distribution. Excluding the 1889 and/or 2009 pandemics changed the median estimated annual infections by up to approximately 13% and increased uncertainty intervals but did not alter the conclusion that several thousand human AIV infections are expected to occur annually. Sensitivity analyses of death reporting showed that lower reporting completeness increased the estimated IFR, although the qualitative conclusions were unchanged. A mechanistic boundary analysis evaluating extreme combinations of key evolutionary and epidemiological parameters produced annual human AIV infection estimates ranging from approximately 1,300 to 21,000 infections per year, corresponding to IFR estimates ranging from approximately 0.10% to 1.60% (S1 Appendix A.3).

## Discussion

We developed a stochastic model based on epidemiological and evolutionary processes that give rise to pandemics to estimate the number of human infections with AIVs that occur each year for the years when a pandemic is not occurring. The median estimated mean annual number of human AIV infections worldwide was 6,441 infections per year. This estimate is much higher than the 986 human cases reported to date and suggests that many infections are undetected and could be because some humans infected with AIVs are asymptomatic or symptomatic but not tested. Based on our estimate of annual AIV infections and the assumption that reported human H5N1-associated deaths provide a reasonable proxy for fatality among AIV infections, the estimated IFR of 32 deaths per 10,000 infections is much lower than the reported H5N1 case fatality rate of 48% [15]. The estimated IFR better informs the risk of AIV infections among people who handle infected wildlife as it includes asymptomatic and undiagnosed individuals [17] and is not biased by the intensity of surveillance activities.

Serological studies among occupationally exposed populations provide independent evidence that human AIV infections are frequently undetected. In two studies conducted by the U.S. Centers for Disease Control and Prevention, H5 antibodies were detected in 7% of dairy farm workers and 2% of bovine veterinarians despite the absence of recognized infection [16,36,37]. As an approximate illustration, if 7% seropositivity occurred among 100,000 exposed workers, this would correspond to approximately 7,000 infections over the relevant exposure period.

Although this calculation cannot be directly compared with our estimated global annual average of approximately 6,000 human AIV infections because the populations, geographic settings, and exposure periods differ, it demonstrates that undetected infections among high-risk groups alone could occur at an order of magnitude consistent with our model estimates. Together, these findings support the conclusion that substantial numbers of zoonotic AIV infections may go unrecognized by routine surveillance.

Our estimated IFR of 32 (95% uncertainty interval: 9.6, 75) deaths per 10,000 infections (0.32%) for humans infected with AIVs is high but associated with substantial uncertainty. For our model that assumes pandemics are becoming more recent, we estimated an IFR in 2026 of 16 (95% uncertainty interval: 5.6, 33) deaths per 10,000 infections. The IFR of SARS-CoV-2 in 2020- 2023 was estimated as 16 deaths per 10,000 infections [34] and 2.9-6.5 deaths per 10,000 infections for seasonal influenza [35]. Therefore, even with the uncertainty in our estimates, both due to the few observations of pandemics that were considered in estimating the IFR and due to uncertainty in whether pandemics are become more recent, our estimate of the IFR for human infections with AIV is similar to, and possibly higher than, the IFR of SARS-CoV-2 during the recent pandemic and higher than that of seasonal human influenza.

Given our estimate of a relatively high IFR for human AIV infections, for hunters, trappers, poultry workers, farmers, wildlife officials, veterinarians and other groups that handle wildlife there is a need to be cautious as there is a relatively high chance that human AIV infections can result in severe infections. The primary virulence mechanism of all H5 viruses involves hemagglutinin (HA), which relies on host proteases for cleavage into two subunits required for cell invasion. In HPAI viruses, a polybasic cleavage site is formed by acquiring multiple basic amino acids. This allows cleavage by furin-like proteases which are distributed more broadly in tissues, enabling systemic infection in birds and mammals, which can result in high pathogenicity [16]. Because the fatality component of our estimate is derived primarily from H5N1 infections, the relatively high estimated IFR is biologically consistent with the virulence mechanisms characteristic of highly pathogenic H5 viruses.

Efforts to prevent human infections with AIVs are necessary given the high estimated individual risk of severe outcomes (as measured by the IFR) and because reducing animal-to-human spillover lowers the risk of pandemic emergence. We estimate that preventing 20% of animal-to-human AIV spillovers annually would delay pandemic emergence by an average of 9.4 years and preventing 50% of spillovers would delay pandemic emergence by 37.5 years. The most commonly reported exposure pathway for human HPAI A(H5N1) virus infection is handling poultry or their secretions [38]. Measures that prevent zoonotic AIV spillover to humans include not touching, feeding or handling potentially infected birds or other animals. When contact cannot be avoided, recommended measures include wearing gloves and a well-fitted respirator or medical mask, reporting infected animals to the appropriate animal health authority [7], the humane destruction of infected and exposed animals, and strict quarantine and animal movement controls to prevent disease spread [39].

We found that the pandemic start date had no clear effect on the mean interpandemic period. The reason that there is not stronger evidence supporting pandemics occurring more frequently now than in the past, is that there are few observations of pandemics (rather than abundant data that support no effect), and because arguments have been presented for mechanisms that would mean pandemics occur more frequently now. These mechanisms include agricultural expansion, deforestation, urbanization and wildlife trade and consumption. These activities bring wildlife, livestock, and humans into close contact which can lead to zoonotic spillovers, epidemics, and pandemics [33]. Increased global connectivity through international travel, population growth, and climate change further increases the risk of infectious disease outbreaks [40].

Given the strong modelling assumptions and limited historical pandemic data underlying the analysis, the sensitivity analyses (S1 Appendix A.3) provide an important assessment of the robustness of our conclusions. Although uncertainty in the probability of a pandemic-capable evolution had the greatest influence on the inferred number of human AIV infections, uncertainty in the historical pandemic data also contributed to variation in the estimates, reflecting the small number of historical pandemics available to estimate the interpandemic distribution. Notably, approximately order-of-magnitude differences in the inferred annual number of infections occurred only under the most extreme plausible parameter combinations, supporting the overall conclusion that a substantial burden of previously undetected human AIV infections is likely.

A limitation of our work is that we inferred the number of human AIV infections from assumed evolutionary mechanisms, parameter value distributions, and the historical rate of pandemics, without validating these estimates using diagnostic testing or serological surveys. Consequently, the validity of our estimates depends on our model assumptions and parameter distributions. Our modelling assumes that the probability of a pandemic-capable evolutionary change occurring is constant over time, but this is likely not the case. Adaption of AIV strains to spread between humans might occur due to reassortment that occurs when a human is coinfected with AIV and human influenza viruses, but this risk of coinfection is variable. The severity of a future AIV pandemic is uncertain as recent human infections with H5N1 clade 2.3.4.4b viruses have had much lower-case fatality ratios than in the past with 2 deaths in 71 cases in the US compared with previous outbreaks in Asia where the case fatality ratio was approximately 50% [6,16].

However, it is unclear whether the recent H5N1 virus infections in humans cause less severe disease or mild cases are merely under-detected in Asia [8].

A further limitation is that the fatality component of our IFR estimate is derived primarily from historical H5N1-associated deaths. Because H5N1 accounts for most documented fatal human AIV infections, we used these deaths as a proxy for fatality among zoonotic AIVs with pandemic potential. Consequently, the estimated IFR should be interpreted as an approximation for this broader class of viruses, informed primarily by H5N1-associated fatality, rather than a subtype-specific H5N1 IFR.

Our approach to estimate the annual number of human AIV infections does not consider the seasonal and wildlife epidemiologic dynamics of zoonotic spillovers or the seasonality of human influenza and the probability of a pandemic-capable mutation occurring. Instead, we assume only seasonal averages for these values, which could result in biased estimation due Jensen’s inequality. In particular, because the transmission term 1/(1 − *R*_0_) is convex, averaging seasonal variation in *R*_0_ before estimating infections will tend to underestimate the expected number of infections, as periods of higher transmissibility contribute disproportionately to total transmission. The magnitude of this bias is uncertain; however, we expect it to be smaller than the uncertainty associated with stochastic variation in pandemic emergence and historical interpandemic periods. We also do not consider the spatial components of AIV spread, such that our estimates are for the number of human AIV infections that occur worldwide, and our modelling approach is not sufficient to inform where these infections might be occurring in any given year, or how these infections could change between years. Future directions of this work include adding temporal and spatial structure to the model to better inform when, and where, our inferred number of human AIV infections are occurring.

Given the recent outbreaks of HPAI A(H5N1) virus in mammals, it is necessary to communicate the risk to humans and to explain the rationale for preventative measures. One component of risk to hunters, trappers, poultry workers, farmers, wildlife officials, veterinarians and other groups that handle wildlife is the chance that they are handling an infected animal. Initiatives such as AviFluMap and similar projects in the United States and Europe [41], aim to estimate this component of risk. Another component of risk is the probability of a severe outcome following a human AIV infection, and estimating this quantity was the focus of our study. The strength of our work is that we estimated undetected human AIV infections by combining sources given the limited data available to estimate this quantity. These data are historical records of pandemics, and this approach is reasonable given that there are few serological surveillance activities to detect asymptomatic and other untested human AIV infections. We estimate that the IFR of human AIV infections is high relative to human influenza, and possibly SARS-CoV-2, such that individuals that take measures to prevent AIV infections would avoid the high risk of a severe outcome. We also show that the value of preventing human AIV infections extends beyond the individual, as preventing human AIV infections delays the emergence of a global pandemic. Our results quantify the value of preventing human AIV infections to both the individual and to society.

## Acknowledgements

We acknowledge comments on the manuscript from Andrew Lang, Joseph Baafi, Francis Anokye, Abdou Fofana and Giuseppe Pasqualino.

During the preparation of this manuscript, ChatGPT (OpenAI) was used to assist with editing for grammar and clarity and to provide coding assistance for data analysis and figure preparation. All code, analyses, results, and interpretations were reviewed and verified by the corresponding author, who takes full responsibility for the content of this manuscript.

## Data Availability

All code and output used in the manuscript are available in this repository: https://github.com/jmack14/Estimating_the_IFR_of_zoonotic_AIVs_with_pandemic_potential

## Supporting information

## S1 Appendix

### A.1 Parameter distributions

Fig A1. shows the sampled values of the parameters that were used to estimate the mean annual number of human AIV infections, where *σ* genotypes are created by sampling from plausible trait values.

**Fig A1.**
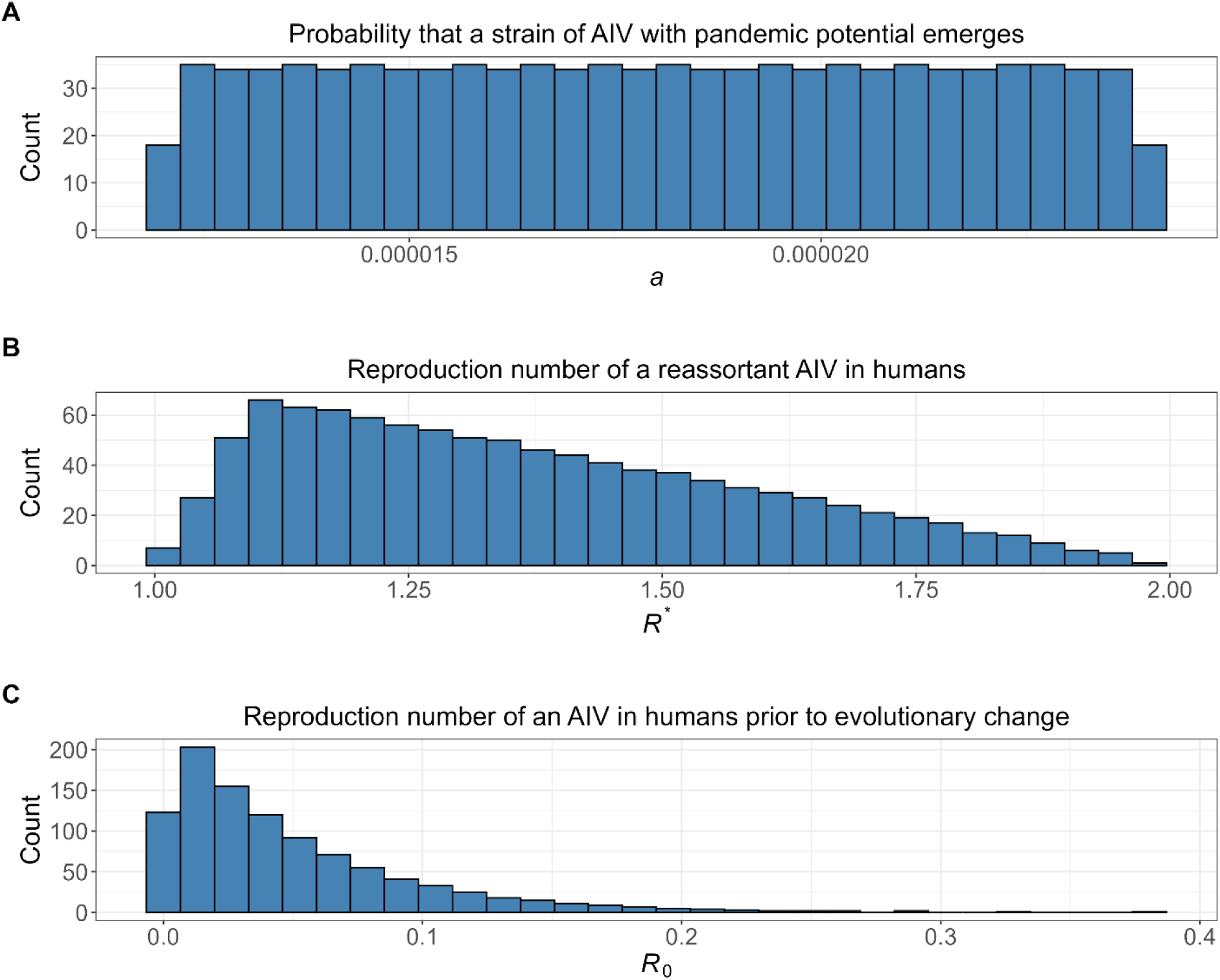
Sampled values of three parameters, where these samples are the basis for the estimated annual number of human AIV infections shown in Fig 2.

### A.2 Distribution of interpandemic periods

Fig A2. shows the fitted gamma distribution to the interpandemic periods of the seven influenza pandemics of zoonotic origin that occurred during the past 245 years (Table 2 and Fig 1) with no effect of recency on the mean interpandemic period.

**Fig A2.**
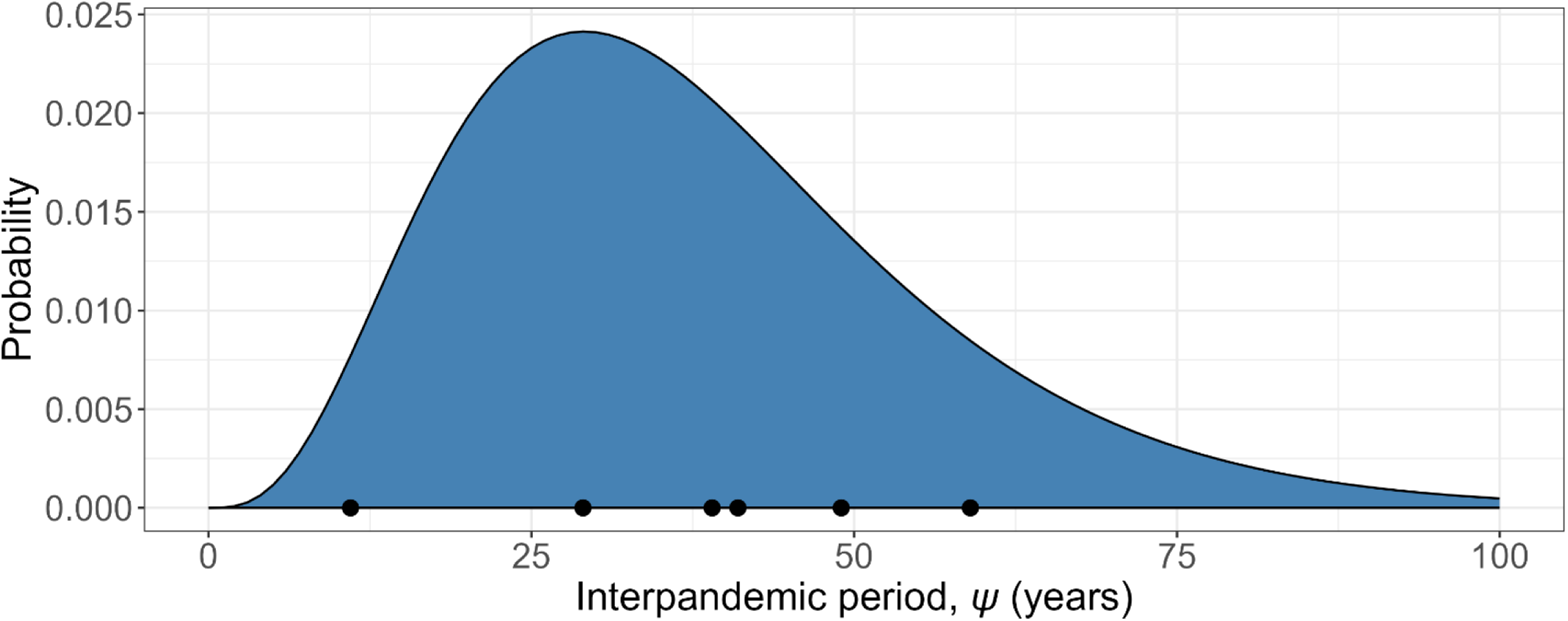
A gamma distribution fitted to observations of the interpandemic periods (black dots) of the seven influenza pandemics that occurred during the last 245 years with no effect of recency on the mean interpandemic period.

For our analysis of the effect of pandemic start date on the mean interpandemic period, we estimated that *δ*_1_ = −0.14 (75% confidence interval: -0.27, 0.029) years per year. The parameter estimate for *δ*1 means that there are approximately 14 years less between pandemics that occur a century later (Fig A3). Based on these estimates, the mean interpandemic period for our earliest record in 1781 was 53.5 years. For the present year, which we define as 2026, the estimated mean interpandemic period was 18.6 years. To investigate the impact of pandemics occurring more recently in recent times, we consider these interpandemic periods for 1781 and 2026 in Fig 2A.

**Fig A3.**
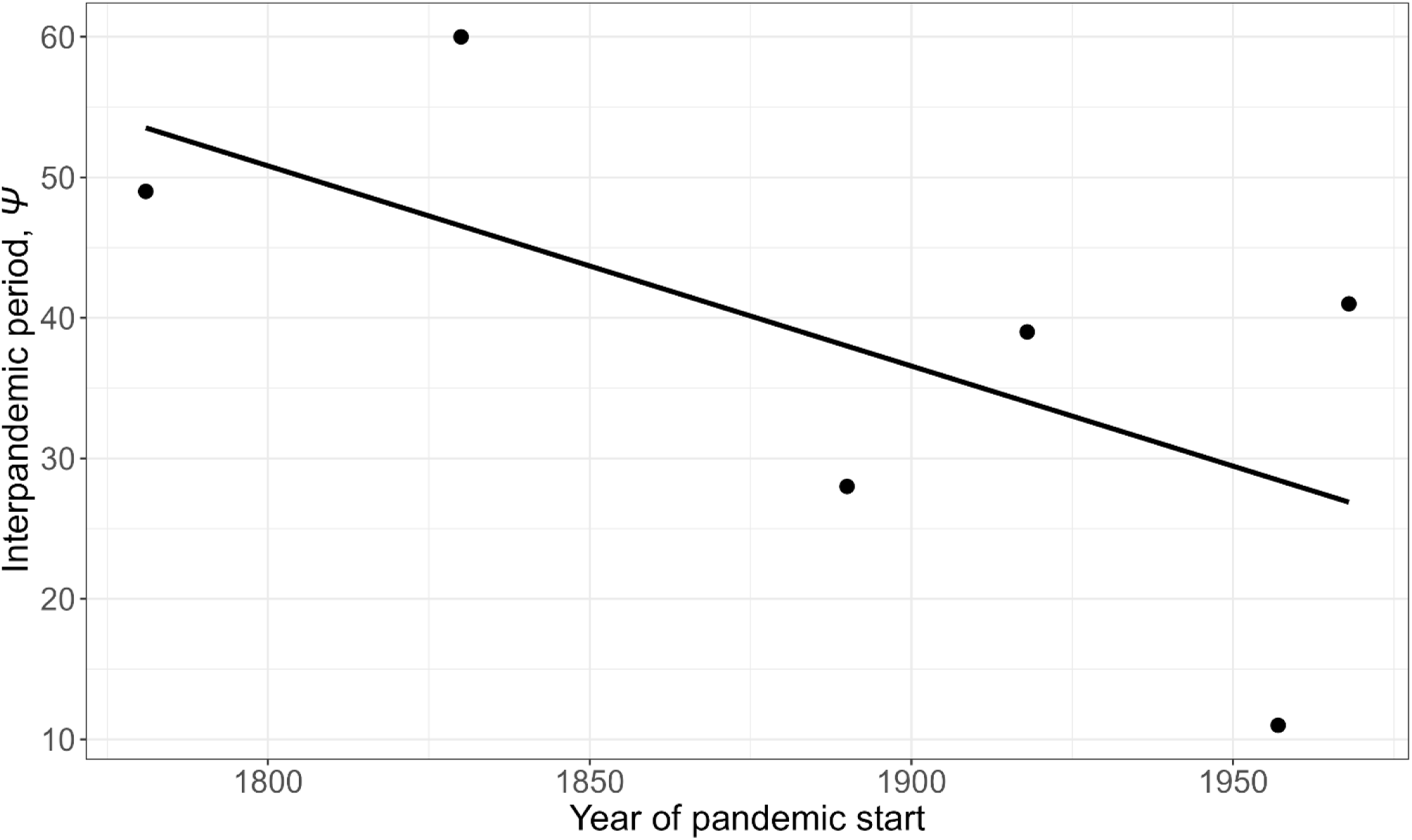
Effect of pandemic start date on the mean interpandemic period.

### A.3 Sensitivity analyses

#### A.3.1 Sensitivity to key model parameters

To evaluate the robustness of the estimated annual number of human AIV infections to assumptions regarding key model parameters, we performed one-way sensitivity analyses. The baseline distributions were described by their distribution parameters. The basic reproduction number, *R*_0_, was sampled from an exponential distribution with a rate parameter *λ_R_* = 20, corresponding to a mean of 1/*λ_R_* = 0.05. The adapted-virus reproduction number, *R*^∗^, was sampled from a triangular distribution with a lower bound of 1, upper bound of 2, and mode parameter *c* = 1.10. The probability of pandemic-capable evolution, *a*, was sampled from a uniform distribution with lower and upper bounds of *a*_min_ = 0.000012 and *a*_max_ = 0.000024.

Sensitivity analyses were performed by varying one distribution parameter at a time while holding all other model assumptions constant. For *R*_0_, the exponential rate parameter was varied from the baseline value of *λ_R_* = 20 to *λ_R_* = 15 and *λ_R_* = 25. For *R*^∗^, the mode parameter was varied from the baseline value of *c* = 1.10 to *c* = 1.05 and *c* = 1.20, while the lower and upper bounds remained fixed. For *a*, the lower and upper bounds of the uniform distribution were simultaneously halved (*a*_min_ = 0.000006, *a*_max_ = 0.000012) and doubled (*a*_min_ = 0.000024, *a*_max_ = 0.000048). All other model assumptions remained unchanged (Table A.1).

**Table A.1.** Sensitivity of estimated annual human AIV infections and IFRs to assumptions regarding key model distribution parameters.

| Scenario | Median estimated mean annual infections | 95% uncertainty interval (infections per year) | Median IFR (%) | 95% uncertainty interval for IFR (%) |
| --- | --- | --- | --- | --- |
| Primary analysis | 6,441 | 2,738–21,160 | 0.32 | 0.096–0.75 |
| $\lambda_R = 15$ | 6,440 | 2,734–21,251 | 0.32 | 0.097–0.75 |
| $\lambda_R = 25$ | 6,428 | 2,722–21,258 | 0.32 | 0.097–0.76 |
| $c = 1.05$ | 6,735 | 2,866–22,127 | 0.31 | 0.093–0.72 |
| $c = 1.20$ | 5,958 | 2,530–19,775 | 0.35 | 0.10–0.81 |
| $a$ range halved | 12,949 | 5,487–43,039 | 0.16 | 0.048–0.36 |
| $a$ range doubled | 3,214 | 1,362–10,602 | 0.64 | 0.19–1.51 |

The estimated annual number of human AIV infections and corresponding IFR were relatively insensitive to plausible changes in the assumed distributions of *R*_0_ and *R*^∗^. In contrast, the estimates were most sensitive to assumptions regarding the distribution of *a*, with lower values of the uniform distribution producing higher inferred infection burdens and lower IFRs, and higher values producing the converse.

#### A.3.2 Sensitivity to genotype weighting

We assessed the sensitivity of the estimated annual number of human AIV infections and the resulting IFRs to assumptions regarding the relative weighting of sampled AIV genotypes. In the primary analysis, all sampled genotypes were assigned equal weights (*ρ_i_* = 1/*σ*) because genotype-specific frequencies among zoonotic AIVs with pandemic potential are not currently known.

To evaluate the influence of this assumption, we considered three alternative genotype weighting schemes while keeping all other model assumptions unchanged. These included (i) genotype frequencies sampled from a Dirichlet distribution with concentration parameter *α* = 1, representing substantial uncertainty in genotype frequencies and allowing highly unequal genotype weights; (ii) weighting proportional to *R*_0_, representing a scenario in which genotypes with greater human transmissibility are assumed to be more common among zoonotic infections; and (iii) weighting proportional to 1 − *R*_0_, representing a scenario in which genotypes with lower human transmissibility are assumed to be more common among zoonotic infections. For each sensitivity scenario, the same sampled genotypes were used, and only the genotype weighting scheme was changed. The estimated annual number of human AIV infections and IFR were recalculated under each alternative weighting scheme and compared with the primary equal-weighting analysis (Table A.2).

**Table A.2.** Sensitivity of estimated annual human AIV infections and IFR to assumptions regarding genotype weighting.

| Scenario | Median estimated mean annual infections | 95% uncertainty interval (infections per year) | Median IFR (%) | 95% uncertainty interval for IFR (%) |
| --- | --- | --- | --- | --- |
| Equal weighting (primary analysis) | 6,441 | 2,738–21,160 | 0.32% | 0.096–0.75 |
| Dirichlet weighting ($\alpha = 1$) | 6,337 | 2,712–21,037 | 0.33% | 0.098–0.76 |
| Weighted proportional to $R_0$ | 6,525 | 2,793–21,660 | 0.32% | 0.095–0.74 |
| Weighted proportional to $1 - R_0$ | 6,447 | 2,759–21,402 | 0.32% | 0.096–0.75 |

Alternative genotype weighting assumptions had little effect on the estimated annual number of human AIV infections or the resulting IFRs. Median annual infection estimates varied from 6,337 to 6,525 infections per year, with all scenarios producing highly overlapping 95% uncertainty intervals. Similarly, median IFR estimates varied only from 0.32% to 0.33%. These results suggest that the assumption of equal genotype weighting is unlikely to be a major source of uncertainty relative to other model assumptions evaluated in this analysis.

#### A.3.3 Sensitivity to the historical pandemic dataset

We assessed the sensitivity of the estimated annual number of human AIV infections and the resulting IFRs to uncertainty in the historical pandemic dataset used to estimate the interpandemic period distribution. The primary analysis included all historical influenza pandemics from 1781 to 2009. However, the virological attribution of the 1889 pandemic remains uncertain [29], and the 2009 H1N1 pandemic originated from swine rather than direct AIV spillover [30].

**Table A.3.** Sensitivity of estimated annual human AIV infections and IFR to alternative historical pandemic datasets.

| Scenario | Median estimated mean annual infections | 95% uncertainty interval (infections per year) | Median IFR (%) | 95% uncertainty interval for IFR (%) |
| --- | --- | --- | --- | --- |
| Primary analysis | 6,441 | 2,738–21,160 | 0.32 | 0.096–0.75 |
| Exclude 1889 pandemic | 5,586 | 1,987–27,284 | 0.37 | 0.076–1.04 |
| Exclude 2009 pandemic | 6,648 | 2,674–24,906 | 0.31 | 0.083–0.77 |
| Exclude both pandemics | 5,624 | 1,808–34,733 | 0.37 | 0.059–1.14 |

To evaluate the influence of these historical uncertainties, we repeated the complete analysis under three alternative pandemic datasets while keeping all other model assumptions unchanged. These included (i) exclusion of the 1889 pandemic, (ii) exclusion of the 2009 pandemic, and (iii) exclusion of both the 1889 and 2009 pandemics (Table A.3).

The estimated annual number of human AIV infections was sensitive to the choice of historical pandemic dataset, although the qualitative conclusions remained unchanged. Excluding the 1889 pandemic reduced the median estimated annual infections by approximately 13%, whereas excluding the 2009 pandemic increased the estimate by approximately 3%. Excluding both events produced a similar reduction to excluding the 1889 pandemic alone and resulted in a wider uncertainty interval, reflecting the reduced amount of historical information available to estimate the interpandemic distribution. Despite these quantitative differences, all scenarios produced estimates on the order of several thousand annual human infections and corresponding IFR estimates of approximately 0.30–0.40%, supporting the overall conclusion that substantial numbers of human AIV infections are likely to occur each year.

#### A.3.4 Sensitivity to reporting completeness of human AIV deaths

To evaluate the assumption that all human AIV deaths were reported, the infection fatality ratio (IFR) was recalculated assuming reporting completeness of 80%, 60%, and 40% (Table A.4).

**Table A.4.** Sensitivity of the estimated infection fatality ratio to assumptions regarding reporting completeness of human AIV deaths.

| Death reporting completeness | Median IFR (%) | 95% uncertainty interval for IFR (%) |
| --- | --- | --- |
| 100% | 0.32 | 0.096–0.75 |
| 80% | 0.40 | 0.12–0.94 |
| 60% | 0.53 | 0.16–1.25 |
| 40% | 0.80 | 0.24–1.87 |

Lower reporting completeness increased the estimated IFR, although the qualitative conclusions remained unchanged.

#### A.3.5 Mechanistic boundary analysis of key model assumptions

To further assess the influence of distributional assumptions on model estimates, we performed a mechanistic boundary analysis in which key model inputs were fixed at extreme values within their assumed ranges. This analysis was designed to evaluate the range of annual human AIV infections and IFR estimates that could arise under combinations of values producing extreme estimates of the mean probability that an AIV infection results in a pandemic, *π̄*, and the mean number of human infections generated per zoonotic spillover, *m̄* .

The boundary values of *π̄* were calculated using Equation (7) by combining extreme values of the probability of pandemic-capable evolution, *a*, the basic reproduction number prior to adaptation, *R*_0_, and the adapted-virus reproduction number, *R*^∗^. For *R*_0_, the lower and upper values were defined as the 2.5th and 97.5th percentiles of the baseline exponential distribution, respectively. For *a*, the minimum and maximum values of the assumed uniform distribution were used. For *R*^∗^, the lower and upper values evaluated in the boundary analysis were used. The minimum value of *π̄* was obtained using the combination of parameter values expected to minimize pandemic establishment probability (low *a*, low *R*_0_, and low *R*^∗^), whereas the maximum value of *π̄* was obtained using the combination expected to maximize pandemic establishment probability (high *a*, high *R*_0_, and high *R*^∗^).

For each resulting value of *π̄*, the mean number of zoonotic spillovers, *n̄_z_*, required to produce the observed annual probability of a pandemic was calculated by solving Equation (6). The corresponding annual number of human infections was then calculated using Equation (4), where *n̄*_*h*_ = *n̄_z_m̄* . The minimum and maximum values of *m̄* were calculated from the extreme values of *R*_0_. The IFR was calculated using the annual number of observed human AIV-associated deaths as the numerator and the boundary estimates of annual human infections as the denominator. Results are presented in Table A.5.

The mechanistic boundary analysis produced annual human infection estimates ranging from approximately 1,300 to 21,000 infections per year, corresponding to IFR estimates ranging from approximately 0.10% to 1.60%. These estimates demonstrate that uncertainty in the evolutionary parameters linking zoonotic infection to pandemic emergence can influence the magnitude of the inferred infection burden. However, even under extreme combinations of parameter values, the analysis continued to support the conclusion that human AIV infections occur substantially more frequently than reported case counts suggest.

**Table A.5.** Mechanistic boundary analysis of key model assumptions.

| Scenario | Value |
| --- | --- |
| Minimum $\bar{\pi}$ | $1.09 \times 10^{-6}$ |
| Maximum $\bar{\pi}$ | $1.47 \times 10^{-5}$ |
| Corresponding $\bar{n}_z$ (lower infection estimate) | 1,282 |
| Corresponding $\bar{n}_z$ (higher infection estimate) | 17,274 |
| Minimum $\bar{m}$ | 1.00 |
| Maximum $\bar{m}$ | 1.23 |
| Minimum annual infections ($\bar{n}_h$) | 1,284 |
| Maximum annual infections ($\bar{n}_h$) | 21,181 |
| Minimum IFR (%) | 0.10 |
| Maximum IFR (%) | 1.60 |

